# Comparative Clinical Outcomes and Mortality in Prisoner and Non-Prisoner Populations Hospitalized with COVID-19: A Cohort from Michigan

**DOI:** 10.1101/2020.08.08.20170787

**Authors:** Ahmed M. Altibi, Bhargava Pallavi, Hassan Liaqat, Alexander A. Slota, Radhika Sheth, Lama Al Jebbawi, Matthew E. George, Allison LeDuc, Enas Abdallah, Luke R. Russel, Saniya Jain, Nariné Shirvanian, Ahmad Masri, Vivek Kak

## Abstract

**Background:** Prisons in the United States have become a hotbed for spreading Covid-19 among incarcerated individuals. Covid-19 cases among prisoners are on the rise, with more than 46,000 confirmed cases to date. However, there is paucity of data addressing clinical outcomes and mortality in prisoners hospitalized with Covid-19.

**Methods:** An observational study of all patients hospitalized with Covid-19 between March 10 and May 10, 2020 at two Henry Ford Health System hospitals in Michigan. Clinical outcomes were compared amongst hospitalized prisoners and non-prisoner patients. The primary outcomes were intubation rates, in-hospital mortality, and 30-day mortality. Multivariable logistic regression and Cox-regression models were used to investigate primary outcomes.

**Results:** Of the 706 hospitalized Covid-19 patients (mean age 66.7 ± 16.1 years, 57% males, and 44% black), 108 were prisoners and 598 were non-prisoners. Compared to non-prisoners, prisoners were more likely to present with fever, tachypnea, hypoxemia, and markedly elevated inflammatory markers. Prisoners were more commonly admitted to the intensive care unit (ICU) (26.9% vs. 18.7%), required vasopressors (24.1% vs. 9.9%), and intubated (25.0% vs. 15.2%). Prisoners had higher unadjusted inpatient mortality (29.6% vs. 20.1%) and 30-day mortality (34.3% vs. 24.6%). In the adjusted models, prisoner status was associated with higher in-hospital death (odds ratio, 1.95; 95% confidence interval (CI), 1.07 to 3.57) and 30-day mortality (hazard ratio, 1.92; 95% CI, 1.24 to 2.98).

**Conclusions:** In this cohort of hospitalized Covid-19 patients, prisoner status was associated with more severe clinical presentation, higher rates of ICU admissions, vasopressors requirement, intubation, inhospital mortality, and 30-day mortality.

## INTRODUCTION

In 2016, there were nearly 2.2 million incarcerated individuals in U.S correctional facilities. Incarcerated individuals are among the most vulnerable for the outspread of infectious diseases due to restricted mobility, overcrowding, confined spaces, limited resources and inequitable access to healthcare services.(1,2) The first known case of Coronavirus Disease 2019 (COVID-19) in a U.S prison occurred in mid-March at the Rikers Island Correctional Facility of New York City. Within few weeks, over 200 cases were diagnosed despite all efforts to curb its spread within the facility.(2) Since then, the outspread of COVID-19 in U.S prisons continued to rampage, with no sign of a respite.(3) As of June 16, 2020, at least 46,249 inmates in U.S. prisons were reported to have tested positive for COVID-19 and 548 died.(3) Nationwide estimates for COVID-19 infection rates among prisoners are five times higher than in general population.(3)(4) Despite this mortifying public health crisis, current federal and state-level healthcare policies failed to respond adequately in a manner that would lead to “flattening the curve” in U.S prisons.(1)

In the absence of an effective vaccine, physical distancing strategies remain the most effective mitigative measure to halt the rate of COVID-19 spread. Such strategies were demonstrated to be effectual during the Spanish influenza pandemic when early, sustained, and sweeping imposition of public gathering restrictions in the 1918 managed to cease the flu’s death toll in cities across America.(5) Yet, in situations where practicing physical distancing has not been attainable, infection rates and mortality statistics can be astounding - as was seen on cruise ships (e.g., *Diamond Princes* cruise ship) and nursing home facilities.(6) Similarly, physical distancing is practically impossible in the setting of incarceration unless major shift in policies occurs. This challenge has led some local authorities in the U.S to adopt depopulation measures to decrease the number of incarcerated individuals in prisons.

The State of Michigan has been a hotbed for COVID-19 spread among prisoners. With a total of 4,000 documented cases, Michigan ranks third across the nation with the number of COVID-19 cases per 10,000 prisoners.(3) The Michigan Department of Correction has adopted a strategy of universal nasopharyngeal swab testing for Severe Acute Respiratory Syndrome Coronavirus 2 (SARS-CoV-2) for all prisoners aiming to identify asymptomatic carriers. By the end of May, 2020, a total of 38,130 prisoners have been tested and 3,982 (10.4%) were found to be positive for SARS-CoV-2.(7) Clinical data about outcomes of prisoners with COVID-19 are lacking. In this study, we sought to report on the characteristics and clinical outcomes of prisoners hospitalized with COVID-19 as compared to non-prisoners.

## METHODS

### 1. Study Design, Settings, and Population

This was a retrospective cohort study of all consecutive patients hospitalized between March 10 and May 10,2020 and tested positive for SARS-CoV-2 on qualitative polymerase chain-reaction (PCR) assay at two Henry Ford Health System hospitals. Patients evaluated at the emergency department without being hospitalized were excluded. The first admission with COVID-19 was considered as the index admission; data was extracted from the index admission while data from readmissions were disregarded. The study was reviewed and approved by the Henry Ford Health System (HFHS) Institutional Review Board (IRB). Patients included in the study were those hospitalized at two Henry Ford hospitals (i) Henry Ford Allegiance Health: a 475-bed hospital located in Jackson county, Michigan and (ii) Henry Ford West Bloomfield: a 200-bed hospital located in Oakland county, Michigan. Admission criteria and treatment protocols were similar at both hospitals based upon healthcare system’s COVID-19 management guidelines. Patients or the public were not involved in the design, conduct, reporting, or dissemination plans of our research.

### 2. Data Collection

We collected patient-level data on imprisonment status (prisoner versus non-prisoner), demographic characteristics (age, sex, and race), chronic medical conditions, smoking status, and obesity (determined by body-mass index). Additional information included patient-reported symptoms, vital signs, medications, laboratory findings, complications, and treatment outcomes. Reported clinical data were linked to inpatient encounters during which a patient tested positive for COVID-19, or first inpatient encounters that followed an out-of-hospital positive COVID-19 test. Upon hospitalization, COVID-19 testing was repeated to confirm diagnosis when initial positive COVID-19 testing could not be documented. Follow-up information was obtained by contacting patients, families, or nursing home facilities via phone. Prisoners’ follow-up information was obtained by contacting the healthcare staff at the prison facilities.

### 3. Statistical Analysis

Characteristics of COVID-19 patients were compared according to imprisonment status (prisoners vs. non-prisoners). For continuous variables, measures are presented as means and standard deviations (SD) or medians and interquartile ranges (IQRs). Categorical measures are presented as percentages. The prespecified primary outcomes were intubation, in-hospital mortality, and 30-day mortality. Each outcome measure was assessed using unadjusted and adjusted models. Factors associated with in-patient mortality and intubation were examined with the use of multivariable logistic regression models. Factors associated with 30-day mortality were investigated with the Cox proportional-hazards models. Each of the primary outcomes was examined using four models: (i) model 1: included imprisonment status only, (ii) model 2: included imprisonment status, age, and sex, (iii) model 3: included model 2 variables, obesity, and Charlson Comorbidity Index (CCI) score, (iv) model 4: included model 3 variables, respiratory rate, lymphocyte count, platelets, creatinine, aspartate aminotransferase (AST), lactate dehydrogenase (LDH), c-reactive protein (CRP), ferritin, procalcitonin, and d-dimer upon admission. CCI score was calculated on the basis of 17 comorbid conditions, each of which is assigned a weighted score.(8) A higher score on the CCI indicates a greater comorbidity burden. All model covariates were selected a priori based on clinical relevance, demonstrated association with clinical outcomes in previous studies, or bivariate analyses with outcomes.

Data for intubation and in-hospital mortality were complete for the whole cohort; while data for 30-day mortality was missing for 9 patients, all from the non-prisoner group. Patients with missing 30-day mortality were not included in the Cox proportional-hazards models. Missing data for covariates were handled with the use of multiple imputation. Multiple imputation models incorporated all available baseline data. The following covariates were imputed in both the logistic and Cox regression models: race, lymphocyte count, platelets, LDH, CRP, ferritin, procalcitonin, and d-dimer.

Results from the analyses of inpatient mortality and intubation are reported as odds ratios and results from the analyses of 30-day mortality are reported as hazard ratios. The proportional-hazards assumption for Cox models was investigated based on Schoenfeld residual method as well as graphically. Survival curves were plotted using the Kaplan-Meier method to compare 30-day mortality between the two groups. Log-Rank test was used to test equality of survival functions. A *P* value <0.05 was considered statistically significant. All statistical analyses were performed with the use of STATA 16 (State Corp LLC, College Station, Texas).

## RESULTS

### 1. Characteristics of the Cohort

The first case of COVID-19 among prisoners in Michigan was identified on March 22, 2020; while the first hospitalization for a prisoner in our hospitals was on March 25, 2020. A total of 706 consecutive patients with COVID-19 were included in the analyses. Of them, 108 patients (15.3%) were prisoners and 598 were non-prisoners. The mean age of patients in the cohort was 66.7 ± 16.1 years, 56.5% were males (98.0% of prisoners were males), 44.3% were Black and 48.0% were Caucasians. Overall, prisoners had a higher CCI score (1.85 versus 1.57) and a higher prevalence of COPD, and underlying malignancies. While, non-prisoners were older (67.7 vs. 61.2 years) and had a higher prevalence of chronic kidney disease, obesity, and dementia. Comorbidities in both groups are detailed in table 1.

**Table 1:** Demographics and Baseline Characteristics for Hospitalized COVID-19 Positive Patient, Comparing Prisoners to Non-prisoners.

|  | Prisoners<br>(N= 108) | Non-prisoners<br>(N= 598) |
| --- | --- | --- |
| Male, n. (%) | 106 (98.2%) | 293 (49.0%) |
| **Age, years<sup>a</sup>** | 61.2 ± 12.5 | 67.7 ± 16.5 |
| **Age >65 years** | 40 (37.0%) | 337 (56.4%) |
| **Body Mass Index (BMI), kg/m<sup>2</sup>**<br>Obesity (BMI ≥30)<br>Morbid Obesity (BMI≥40) | 29.5 ± 5.8<br>45 (41.7%)<br>5 (4.6%) | 30.4 ± 7.9<br>276/595 (46.4%)<br>70/595 (11.8%) |
| **Race<sup>a</sup>**<br>Caucasian<br>Black<br>Others | 47 (43.5%)<br>54 (50.0%)<br>7 (6.5%) | 285/583 (48.9%)<br>252/583 (43.2%)<br>46/583 (7.9%) |
| **Smoking Status<sup>a</sup>**<br>Non-smoker<br>Active smoker<br>Former smoker | 19/96 (19.8%)<br>3/96 (3.1%)<br>74/96 (77.1%) | 355/569 (62.4%)<br>25/569 (4.4%)<br>189/569 (33.2%) |
| Hypertension<sup>b</sup> | 76 (70.4%) | 438 (73.2%) |
| Dyslipidemia<sup>b</sup> | 54 (50.0%) | 347 (58.0%) |
| Coronary Artery Disease<sup>b</sup> | 20 (18.5%) | 117 (19.6%) |
| History of Myocardial Infarction<sup>b</sup> | 15 (13.9%) | 52 (8.7%) |
| History of Congestive Heart Failure<sup>b</sup> | 16 (14.8%) | 98 (16.4%) |
| Peripheral Vascular Disease<sup>b</sup> | 9 (8.3%) | 38 (6.4%) |
| Cerebrovascular Disease<sup>b</sup> | 10 (9.3%) | 70 (11.7%) |
| Chronic Liver Disease<sup>b</sup> | 2 (1.9%) | 14 (2.3%) |
| Chronic Kidney Disease<sup>b</sup><br>Dialysis Dependent<br>Kidney Transplant | 11 (10.2%)<br>1 (0.9%)<br>0 (0.0%) | 143 (23.9%)<br>14 (2.3%)<br>6 (1.0%) |
| Type 2 Diabetes Mellitus (DM) <sup>b</sup><br>Insulin-dependent DM<br>Complicated DM* | 39 (36.1%)<br>14 (12.9%)<br>5 (4.6%) | 199 (33.3%)<br>51 (8.5%)<br>68 (11.4%) |
| Malignancy<sup>b</sup><br>Solid Tumors<br>Leukemia/Lymphoma | 8 (7.4%)<br>3 (2.8%) | 15 (2.5%)<br>7 (1.2%) |
| Dementia<sup>b</sup> | 2 (1.9%) | 108 (18.1%) |
| Hemiplegia<sup>b</sup> | 1 (0.9%) | 16 (2.7%) |
| HIV <sup>b,c</sup> | 3 (2.8%) | 3 (0.5%) |
| Hepatitis C<sup>b</sup> | 4 (3.7%) | 6 (1.0%) |
| Chronic Lung Disease<sup>b</sup> |  |  |
| COPD<sup>b,c</sup> | 40 (37.0%) | 63 (10.5%) |
| Asthma | 10 (9.3%) | 65 (10.9%) |
| Sleep Apnea | 6 (5.6%) | 73 (12.2%) |
| Interstitial Lung Disease | 0 (0.0%) | 7 (1.2%) |
| Pulmonary Sarcoidosis | 0 (0.0%) | 5 (0.8%) |
| Oxygen Dependent | 6 (5.6%) | 25 (4.2%) |
| CCI Score | 1.85 ± 2.02 | 1.57 ± 1.79 |

<sup>a</sup> Plus-minus values indicate means ± standard deviation (SD). Data for race and smoking status were missing for 15 and 41 patients, respectively. Race and smoking status were self-reported by the patients.
<sup>b</sup> Absence of diagnoses recorded in the medical record was assumed to mean absence of the conditions.
<sup>c</sup> HIV denotes Human Immunodeficiency Virus; COPD denotes chronic obstructive pulmonary disease.

### 2. Clinical Signs and Laboratory Findings

The median time from self-reported onset of symptoms to admission was 6 (IQR: 3-8) days for prisoners and 5 (IQR: 2-8) days for non-prisoners. Compared to non-prisoners, prisoners had worse clinical signs at presentation, including fever (temperature ≥ 38.0 Celsius), hypoxemia (oxygen saturation <94%), and tachypnea (respiratory rate >24/minute). Further, prisoners had markedly higher levels of inflammatory markers upon admission, including CRP, ferritin, and LDH. Clinical signs and laboratory findings upon admission and during hospital course are detailed in table 2.

**Table 2:** Vital Signs and Laboratory Findings in Patients Hospitalized with COVID-19 (Prisoners vs. Non-Prisoners).

|  | **Prisoners**<br>**(N= 108)** | **Non-prisoners**<br>**(N= 598)** |
| --- | --- | --- |
| **Vital Signs Upon Admission** |  |  |
| **Respiratory rate >24** | 48 (44.4%) | 167 (27.9%) |
| **Temperature ≥ 38.0 Celsius** | 29 (26.9%) | 89 (14.9%) |
| **Oxygen saturation <94%** | 52 (48.2%) | 223 (37.3%) |
| **Systolic blood pressure <90 mmHg** | 1 (0.93%) | 27 (4.7%) |
| **Mean arterial pressure (mmHg) <sup>b</sup>** | 89.3 ± 16.7 | 90.1 ± 15.1 |
| **Laboratory Findings Upon Admission** |  |  |
| **White blood cell count <sup>a b</sup>** | 7.2 (5.4 – 9.2) | 6.5 (4.9 – 9.2) |
| **Lymphocyte count <1,000/ul** | 61 (56.5%) | 337/594 (56.7%) |
| **Platelet count <150,000/ul** | 17 (15.7%) | 148/594 (24.9%) |
| **AST >40 U/liter<sup>c</sup>** | 62/106 (58.5%) | 271/572 (47.4%) |
| **ALT >40 U/liter<sup>c</sup>** | 30/107 (28.0%) | 153/572 (26.8%) |
| **Creatinine (mg/dl)<sup>a</sup>** | 1.6 ± 1.9 | 1.8 ± 6.4 |
| **Creatinine >1.5 mg/dl** | 26 (24.1%) | 160/592 (27.0%) |
| **Lactate dehydrogenase <sup>a b</sup> (U/L)** | 412 (322-547) | 288 (218-383) |
| **HS-Troponin I >18 ng/L <sup>c</sup>** | 38 (35.2%) | 212/576 (36.8%) |
| **Procalcitonin (ng/ml)** |  |  |
| <0.25 | 61/106 (57.6%) | 361/532 (67.9%) |
| 0.25 – 0.5 | 24/106 (22.6%) | 75/532 (14.1%) |
| >0.5 | 21/106 (19.8%) | 96/532 (18.0%) |
| **C-Reactive Protein (mg/dl)** |  |  |
| < 5 | 26/107 (24.3%) | 155/559 (27.7%) |
| 5 – 10 | 27/107 (25.2%) | 171/559 (30.6%) |
| >10 | 54/107 (50.5%) | 233/559 (41.7%) |
| **Ferritin (ng/ml)** |  |  |
| >300 | 15/107 (14.0%) | 181/568 (31.9%) |
| 300 – 2,000 | 70/107 (65.4%) | 346/568 (60.9%) |
| ≥ 2,000 | 22/107 (20.6%) | 41/568 (7.2%) |
| **D-dimer (ug/ml)** |  |  |
| < 0.25 | 13/105 (12.4%) | 58/554 (10.5%) |
| 0.25 – 0.5 | 30/105 (28.6%) | 154/554 (27.8%) |
| > 0.5 | 62/105 (59.0%) | 342/554 (61.7%) |
| **Laboratory Findings During Hospital Course** |  |  |
| **Lymphocyte nadir <1,000/ul** | 94 (87.0%) | 492/594 (82.8%) |
| **Platelet nadir <150,000/ul** | 33 (30.6%) | 215/594 (36.2%) |
| **ALT Peak >40 U/liter** | 68/107 (63.6%) | 291/571 (51.0%) |
| **AST Peak >40 U/liter** | 74/106 (69.8%) | 361/572 (63.1%) |
| **HS-Troponin I Peak >18 ng/L** | 48 (44.4%) | 262/576 (45.5%) |
| **Procalcitonin Peak (ng/ml)** |  |  |
| <0.25 | 53/106 (50%) | 340/532 (63.9%) |
| 0.25 – 0.5 | 23/106 (21.7%) | 71/532 (13.4%) |
| >0.5 | 30/106 (28.3%) | 121/532 (22.7%) |
| **C-Reactive Protein Peak (mg/dl)** |  |  |
| < 5 | 16/106 (15.1%) | 111/559 (19.9%) |
| 5 – 10 | 28/106 (26.4%) | 128/559 (22.9%) |
| >10 | 62/106 (58.5%) | 320/559 (57.3%) |
| **Ferritin Peak (ng/ml)** |  |  |
| >300 ng/ml | 8/107 (7.5%) | 127/568 (22.4%) |
| 300 – 2,000 ng/ml | 72/107 (67.3%) | 369/568 (64.9%) |
| ≥ 2,000 ng/ml | 27/107 (25.2%) | 72/568 (12.7%) |
| **D-dimer Peak (ug/ml)** |  |  |
| < 0.25 | 8/103 (7.8%) | 32/554 (5.8%) |
| 0.25 – 0.5 | 24/103 (23.3%) | 97/554 (17.5%) |
| > 0.5 | 71/103 (68.9%) | 425/554 (76.7%) |

<sup>a</sup> Data for white blood cell (WBC) count, creatinine, and lactate dehydrogenase (LDH) upon admission were missing for 4, 6 and 35 patients in the non-prisoner group, respectively.
<sup>b</sup> Plus-minus values indicate means ± standard deviation (SD). Values for WBC count and LDH are presented as median (interquartile range).
<sup>c</sup> AST indicates aspartate aminotransferase; ALT indicates alanine transaminase; HS-troponin indicates high-sensitivity troponin.

### 3. Medical Treatment and Hospital Course

The median length of hospital stay was 8 (IQR: 5-12) days. A total of 141 of 708 patients (19.9%) required admission to the intensive care unit (ICU); 26.9% of prisoners and 18.7% of non-prisoners. During hospital course, 25.6% of patients did not require oxygen support, 41.6% required oxygen support delivered via conventional nasal cannula, 16.4% required high-flow nasal cannula, and 19.9% required intubation. Treatment protocol included anticoagulation (84.0%), corticosteroids (79.4%), antibiotics (46.2%) for prevention/treatment of secondary infections, and hydroxychloroquine (64.8%). Subcutaneous enoxaparin was the anticoagulant of choice in most patients (73%). Corticosteroids were in the form of intravenous methylprednisolone (1-2 mg/kg/day). Tocilizumab, an interleukin-6 inhibitor, was given to 7.2% of patients at a dose of 400 mg intravenously based on clinic criteria. Other treatment modalities include Remdesivir (compassionate use protocol) and Convalescent Plasma (part of Mayo Clinic’s Expanded Access to Convalescent Plasma trial).(9) Of notice, more patients in the prisoners group required pressure support with vasopressors to non-prisoners (24.1% vs. 9.9%). Details of treatment modalities are shown in Table 3. Complications accompanying hospital course are shown in eTable 1 (Supplement 1).

**Table 3:** Treatment, Complications, and Outcomes in Patients Hospitalized with COVID-19 (Prisoners vs. Non-Prisoners).

|  | **Prisoners**<br>**(N= 108)** | **Non-prisoners**<br>**(N= 598)** |
| --- | --- | --- |
| **Therapeutics and Medications** |  |  |
| **Highest O2 Requirement** |  |  |
| Ambient Air | 16 (14.8%) | 150 (25.1%) |
| Nasal Cannula | 23 (21.3%) | 271 (45.3%) |
| High-flow nasal cannula | 38 (35.2%) | 78 (13.0%) |
| NIVM<sup>a</sup> | 4 (3.7%) | 7 (1.2%) |
| IMV<sup>a</sup> | 27 (25.0%) | 92 (15.4%) |
| **Antibiotics** | 55 (50.9%) | 271 (45.3%) |
| **Corticosteroids** | 95 (88.0%) | 466 (78.0%) |
| **Anticoagulation** | 103 (95.4%) | 490 (81.9%) |
| **Hydroxychloroquine** | 49 (45.4%) | 409 (68.4%) |
| **Tocilizumab** | 17 (15.7%) | 34 (5.7%) |
| **Remdesivir** | 2 (1.9%) | 3 (0.5%) |
| **Convalescent Plasma** | 9 (8.3%) | 7 (1.2%) |
| **Renal replacement therapy** | 6 (5.6%) | 24 (4.0%) |
| **Vasopressors** | 26 (24.1%) | 59 (9.9%) |
| **ECMO<sup>a</sup>** | 0 (0.0%) | 2 (0.3%) |
| **Clinical Outcomes** |  |  |
| **Length of Stay (days)** | 9 (5-14) | 7 (5-12) |
| **ICU Admission<sup>a</sup>** | 29 (26.9%) | 112 (18.7%) |
| **Intubation** | 27 (25.0%) | 91 (15.2%) |
| **Inpatient Mortality** | 32 (29.6%) | 120 (20.1%) |
| **14-Day Mortality<sup>b</sup>** | 21 (19.4%) | 102 (17.1%) |
| **30-Day Mortality<sup>b</sup>** | 37 (34.3%) | 145 (24.6%) |
| **30-Day Readmission<sup>b</sup>** |  |  |
| Readmitted | 7/76 (9.2%) | 65/478 (13.6%) |
| Not readmitted | 69/76 (90.8%) | 387/478 (81.0%) |
| Unknown | 0/76 (0.0%) | 26/478 (5.4%) |
| **Disposition Status<sup>c</sup>** |  |  |
| Home | -- | 363 (75.8%) |
| Other Facilities | -- | 96 (20.0%) |
| Hospice | -- | 20 (4.2%) |

<sup>a</sup> NIVM indicates non-invasive mechanical ventilation; IMV indicates invasive mechanical ventilation; ECMO indicates extracorporeal mechanical oxygenation; ICU indicates intensive care unit.
<sup>b</sup> 14-day mortality and 30-day mortality data were missing for 10 patients from the non-prisoner group. 30-day readmission data was missing for 26 patients from the non-prisoner group. Only patients who were discharged alive were included in calculating the 30-day readmission rate.
<sup>c</sup> Disposition status percentages were calculated from the total number of patients discharged alive (479 in the non-prisoner group). Other facilities include skilled nursing facilities and rehabilitation facilities (short-term, long-term, and inpatient rehabilitation).

### 4. Intubation

Intubation and mechanical ventilation during the hospital course was required in 27 of 108 prisoners (25.0%) and 91 of 598 non-prisoners (15.2%). Table 4 shows the unadjusted and adjusted odds ratios of intubation during hospital course. In the crude, unadjusted analysis, prisoners had higher odds of intubation compared to non-prisoners (odds ratio 1.86; 95% CI, 1.14 to 3.03; *P*= 0.01). In the multivariable logistic regression models, the odds ratios of intubation were 1.66 (95% CI, 0.96 to 2.86; *P*= 0.07) and 1.16 (95% CI, 0.63 to 2.14; *P*= 0.64) in models 3 and 4, respectively (eTable 2, Supplement 1).

**Table 4:**
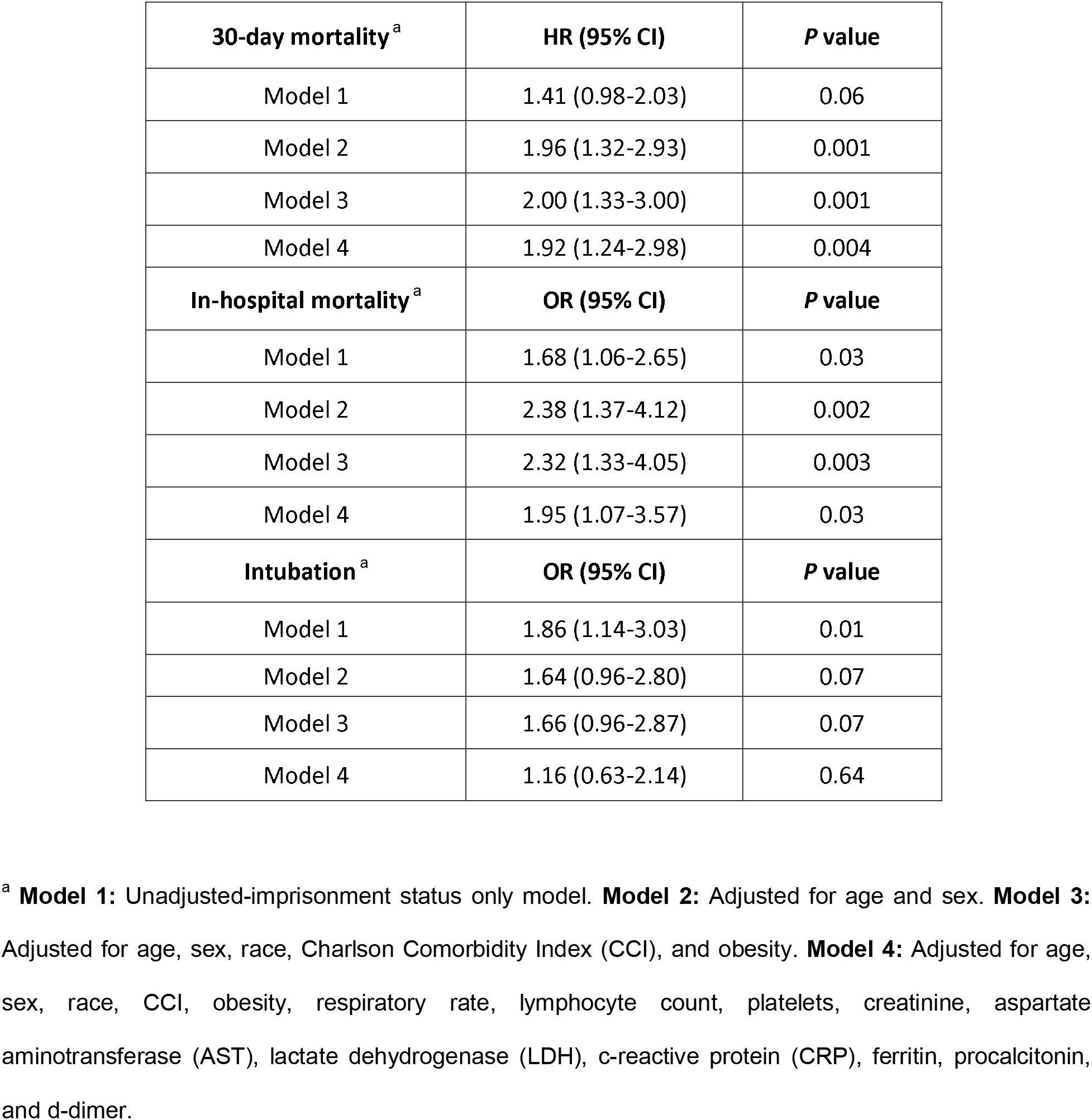
Clinical Outcomes (30-day mortality, in-hospital mortality, and intubation) for Hospitalized COVID-19 Positive Prisoners Compared to Non-prisoners in the Crude Analysis (Model 1) and Multivariable Analyses (Models 2, 3, and 4).

| **30-day mortality <sup>a</sup>** | **HR (95% CI)** | **P value** |
| --- | --- | --- |
| Model 1 | 1.41 (0.98-2.03) | 0.06 |
| Model 2 | 1.96 (1.32-2.93) | 0.001 |
| Model 3 | 2.00 (1.33-3.00) | 0.001 |
| Model 4 | 1.92 (1.24-2.98) | 0.004 |
| **In-hospital mortality <sup>a</sup>** | **OR (95% CI)** | **P value** |
| Model 1 | 1.68 (1.06-2.65) | 0.03 |
| Model 2 | 2.38 (1.37-4.12) | 0.002 |
| Model 3 | 2.32 (1.33-4.05) | 0.003 |
| Model 4 | 1.95 (1.07-3.57) | 0.03 |
| **Intubation <sup>a</sup>** | **OR (95% CI)** | **P value** |
| Model 1 | 1.86 (1.14-3.03) | 0.01 |
| Model 2 | 1.64 (0.96-2.80) | 0.07 |
| Model 3 | 1.66 (0.96-2.87) | 0.07 |
| Model 4 | 1.16 (0.63-2.14) | 0.64 |

<sup>a</sup> **Model 1:** Unadjusted-imprisonment status only model. **Model 2:** Adjusted for age and sex. **Model 3:** Adjusted for age, sex, race, Charlson Comorbidity Index (CCI), and obesity. **Model 4:** Adjusted for age, sex, race, CCI, obesity, respiratory rate, lymphocyte count, platelets, creatinine, aspartate aminotransferase (AST), lactate dehydrogenase (LDH), c-reactive protein (CRP), ferritin, procalcitonin, and d-dimer.

### 5. In-Hospital Mortality

In-hospital all-cause death occurred in 32 of 108 prisoners (29.6%) and in 120 of 598 (20.1%) non-prisoner patients. Mean time from admission to in-hospital death was 12.9 ± 6.6 days for prisoners and 10.4 ± 7.7 days for non-prisoners. Table 4 shows the unadjusted and adjusted odds ratios of in-hospital mortality. In the crude analysis, prisoners had higher odds for in-hospital mortality compared to non-prisoners (odds ratio, 1.68; 95% CI, 1.06 to 2.65; *P*= 0.03). Similarly, in the multivariable logistic regression models (model 4), prisoners had approximately twice the odds of in-hospital death compared to non-prisoners (odds ratio 1.95; 95% CI, 1.07 to 3.57; *P*= 0.03). Variables associated with higher inhospital mortality (etable 3 in Supplement 1) were increasing age, male sex, Caucasian race, increasing CCI, elevated respiratory rate (>24/minute), elevated levels of lactate dehydrogenase (>250 u/L), and elevated procalcitonin levels (>0.25 ng/ml). Imprisonment status was independently associated with increased risk of in-hospital death (model 1).

### 6. 30-Day Mortality

Death within 30-day of follow-up occurred in 37 of 108 prisoners (34.3%) and 145 of 589 non-prisoner patients (24.6%). Mean time from admission to death was 13.7 ± 6.6 days for prisoners and 11.2 ± 7.5 for non-prisoners. Table 4 shows the unadjusted and adjusted hazard ratios of 30-day mortality. In the unadjusted analysis, the hazard ratio of all-cause 30-day mortality was higher for prisoners compared to non-prisoners (hazard ratio 1.41; 95% CI, 0.98 to 2.03; *P*= 0.06). In the adjusted time-to-event analyses (model 4), prisoners had approximately twice the hazard of 30-day mortality compared to non-prisoners (hazard ratio 1.92; 95% CI, 1.24 to 2.98; *P*= 0.004). Variables associated with higher 30-day mortality (etable 4 in Supplement 1) were increasing age, male sex, Caucasian race, increased CCI, and increased respiratory rate (>24/minute). Figure 1 shows the Kaplan-Meier survival curve for mortality from admission to day 30 of follow-up (log-rank test, *P*= 0.06).

**Figure 1:**
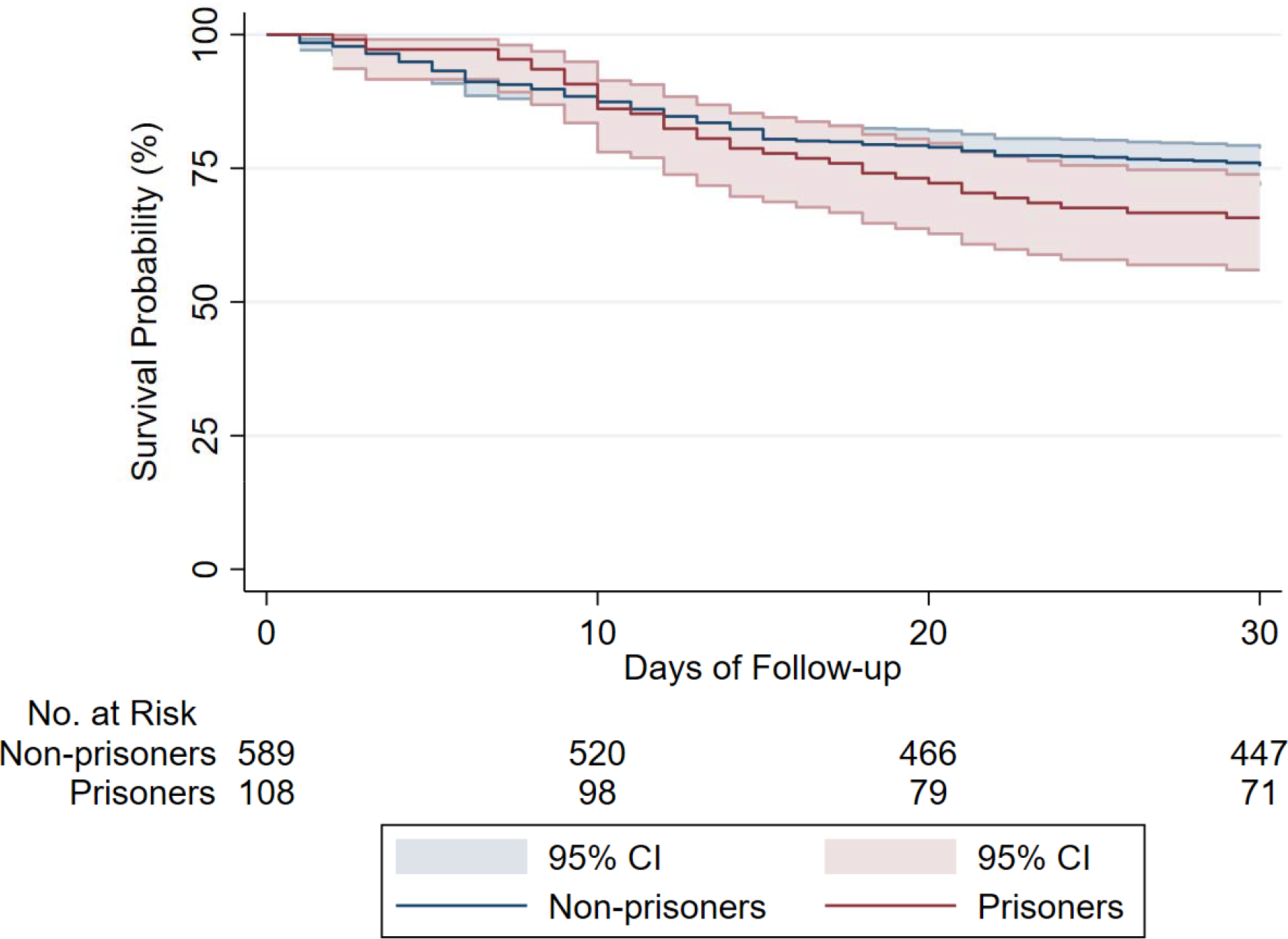
Survival Curve for Hospitalized COVID-19 Positive Patients, by Imprisonment Status. Survival curve shows all-cause mortality for prisoners and non-prisoners (n= 697) hospitalized for COVID-19. Graph is based on a Cox proportional hazards model. Sample size provided reflects 30-day follow-up data availability; data for 30-day mortality was missing for 9 patients from the non-prisoner group. Shaded Area Represent Pointwise 95% Confidence Interval. Log-Rank test for the survival curve had *P* value of 0.06.

The 30-day all-cause mortality for patients requiring intubation was 59.7% (71 of 119 intubated patients died). However, there was a differential mortality based on imprisonment status amongst intubated patients. The 30-day mortality was 88.9% for intubated prisoners compared to 51.1% for non-prisoners. Comparing intubated prisoners to non-prisoners, the unadjusted hazard ratio for 30-day mortality was 2.65 (95% CI, 1.61 – 4.40; *P* <0.001), while the age and gender-adjusted hazard ratio was 2.70 (95% CI, 1.54 – 4.68; *P* <0.001). Kaplan-Meier survival curve at 30-day follow-up among intubated patients (log-rank test, *P*<0.001) is shown in Figure 2.

**Figure 2:**
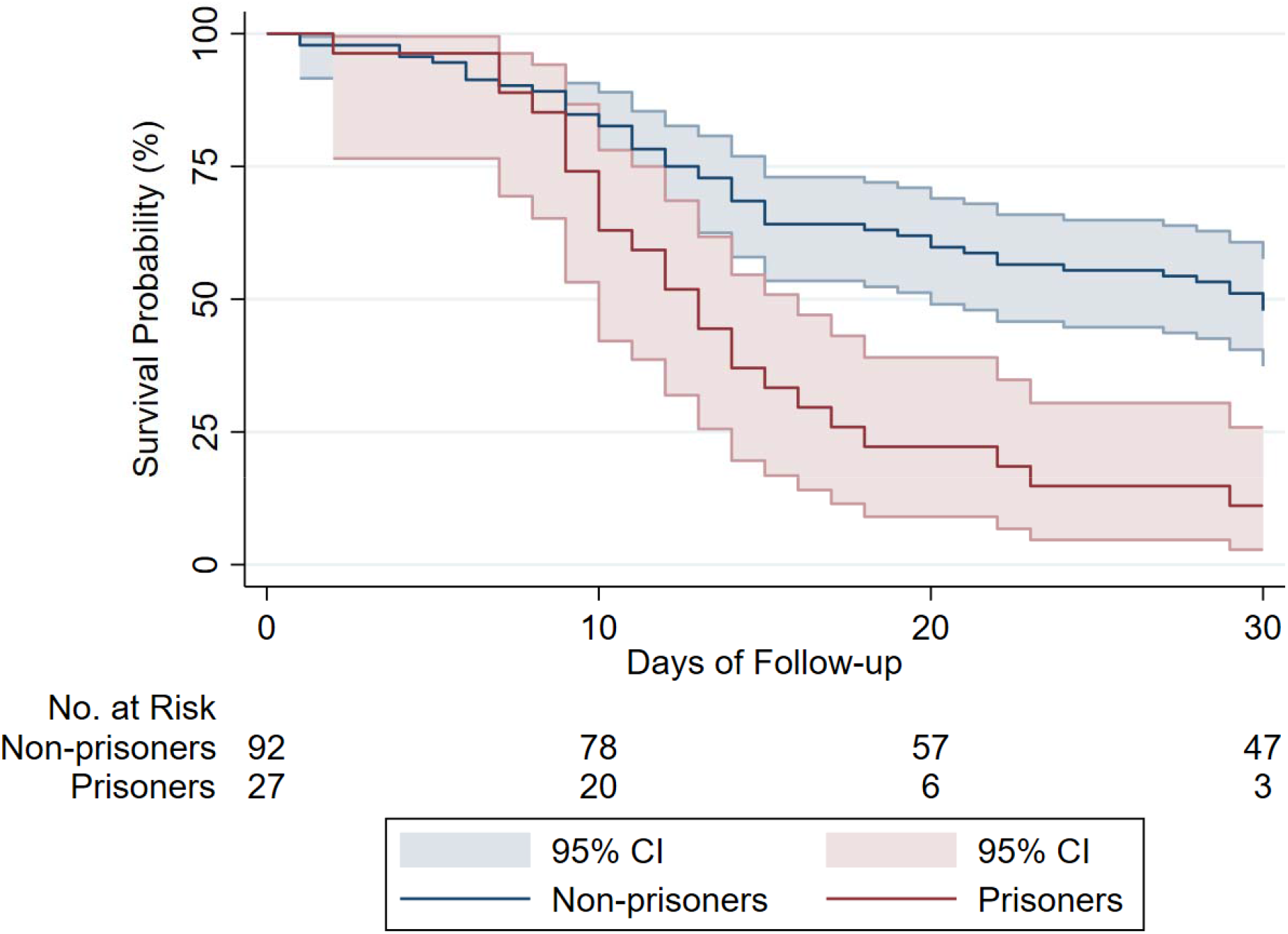
Survival Curve for Intubated COVID-19 Patients, by Imprisonment Status. Survival curve shows all-cause mortality for prisoners and non-prisoners requiring intubation (n= 119). Graph is based on a Cox proportional hazards model. Data on intubation status was complete for all study participants. Data for 30-day mortality was complete for all intubated patients. Shaded Area Represent Pointwise 95% Confidence Interval. Log-Rank test for the survival curve had *P* value of <0.001.

### 7. Disposition at Discharge

For non-prisoners discharged alive (479 patients), 75.8% were discharged to home, 20.0% discharged to other facilities (skilled nursing or rehabilitation facilities), and 4.2% discharged to hospice homes. For prisoners discharged alive (76 patients), clinically stable prisoners were discharged to quarantine units within the prison facility (31.6%), while those requiring additional medical care (i.e., high oxygen requirement) were discharged to a prisoner-designated hospital (68.4%), operated by Michigan Department of Correction.

## DISCUSSION

Despite more than 46,000 reported cases in the U.S, no reports to-date have investigated clinical outcomes and mortality trends among prisoners hospitalized with COVID-19. In this study, we report our experience on the comparative outcomes of 108 prisoners and 698 non-prisoners hospitalized with COVID-19. Prisoner status was associated with approximately twice the risk of death compared to non-prisoners. Prisoners in our cohort presented from four local prisoner facilities located in Jackson and Coldwater City in Michigan. In these facilities, positivity for SARS-CoV-2 was reported to range between 38% (708 of 1,872 at one facility) and 55% (793 of 1,430 at another facility).(7) However, precise estimation for hospitalization rates among COVID-19 positive prisoners was not feasible.

In our cohort, prisoners were more likely to present with signs consistent with Systemic Inflammatory Response Syndrome (SIRS) and hypoxemia. Further, inflammatory markers at presentation (e.g., CRP, ferritin, and LDH) were markedly higher in patients who are prisoners. The higher clinical severity among prisoners could reflect a late reporting of symptoms or a higher threshold for referral of COVID-19 patients from the prison facility to the hospital than that from the community. The Michigan Department of Correction have adopted a pre-hospital scoring system to determine referral decisions.(10) The scoring system is based on age, vital signs, oxygen requirements, and mental status (eTable 5 in Supplement 1); only prisoners meeting the criteria of the scoring system were considered for transportation to the hospital. On the other hand, the need for supplemental oxygen was the primary deciding factor in admitting COVID-19 patients presenting from the community.

A higher percentage of prisoners than non-prisoners required admission to the ICU, pressure support with vasopressors, and intubation. For both in-hospital and 30-day mortality, prisoners had approximately 10% excess mortality rate than non-prisoners. Prisoners who required intubation had strikingly dismal outcomes with 30-day mortality approaching 89%. The discrepancy in mortality rates observed in the study population is multifactorial; while some are attributed to the demographic characteristics and underlying comorbidity burden, others are inherently unique to the imprisonment status. Incarcerated individuals bear a disproportionally higher burden of infectious diseases. Reports have estimated that 25% of all HIV/AIDS cases and 35% of all hepatitis C virus cases in the U.S are among incarcerated individuals.(11) Further, sepsis mortality rates were shown to be four times higher among prisoners to that for non-prisoners (42.5% vs. 15.3%).(12)

In our cohort, prisoners were younger in age, predominantly males, and had higher prevalence of COPD, diabetes mellitus, underlying malignancies, and a higher CCI score. COPD was overrepresented among patients included in this cohort (14.6%) as compared to a reported prevalence of 2 – 6% in large cohorts from New York, Louisiana, and China.(13)(14)(15) Moreover, a disproportionally larger percentage of prisoners had underlying COPD compared to non-prisoners (37.5 vs. 10.5%) – reflecting the higher prevalence of smoking in this group (80.1% were current or former smokers). Data on the association between COPD and disease severity in COVID-19 patients remains scarce. A meta-analysis of limited sample size suggests that COPD patients are at a substantially greater risk of more severe disease and mortality.(16) Nonetheless, the discrepancy in 30-day mortality in our cohort remained remarkable when comparing prisoner to non-prisoner patients without an underlying COPD (29.4% and 23.2%, respectively).

The overwhelming majority of prisoners in our cohorts were males (98.2%). Sex-disaggregated epidemiologic data across 38 countries have shown a significantly higher case-fatality rates for COVID-19 among males.(17) Females have stronger innate, humoral, and cellular immunity resulting in sexspecific outcomes from viral infections.(18)(19) In animal models of mice infected with SARS-CoV-1, higher susceptibility to SARS-CoV-1 infection and greater infiltration of neutrophils and macrophages into the lungs were observed in male mice.(20) Further, ovariectomized and estrogen receptor antagonist-treated female mice had increased mortality from SARS-CoV-1 infection.(20) These findings of sex-based differences in immune response and susceptibility can partially explain the excess mortality from COVID-19 among prisoners.

The study has several limitations. First, the observational study design makes selection bias inevitable, especially considering the potential discrepancy in hospital referral and admission threshold between the two study groups. However, our findings remained largely consistent after controlling for potential confounders related to demographics, comorbidities, clinical signs, and markers of disease severity. Second, the study population was limited to one healthcare system in Michigan admitting patients from certain prisons and communities, and therefore external generalizability requires validation. Finally, presented findings were based on data extracted from the electronic medical records of the health system, and hence, is subjected to the accuracy of documentation by the healthcare team, but the outcomes were adjudicated and all patients discharged alive were contacted directly or through their healthcare office.

In conclusion, our study provides insight into the disparity in clinical outcomes among prisoners hospitalized with COVID-19. With COVID-19 becoming one of the leading causes of death in the U.S., findings on the association between imprisonment and COVID-19 mortality are alarming. Responding to the outbreaks in incarceration facilities is uniquely challenging and failure to mount an adequate response to ameliorate COVID-19 outbreaks in U.S prions has the potential to compromise the well-being of prisoners, correctional workforce, and people living in the communities in which prisons are located. Since physical distancing, the most effective mitigative measure, is constantly violated by prisons overcrowding, major shift in policy towards enforcing ‘mass testing and segregation’ and prisons depopulation (i.e., drastically reduce the population of prisoners) might be warranted to abort further imminent outbreaks.(1) Nonetheless, maintaining the rights of prisoners for timely, adequate, and equitable healthcare services is indispensable to close the gap in COVID-19 clinical outcomes with the general population.

## Data Availability

Data pertaining to this research is available and can be provided upon request from the first author: Dr. Ahmed Altibi

## DISCLOSURE

All authors have completed the ICMJE uniform disclosure form at www.icmje.org/coi_disclosure.pdf and declare: no support from any organization for the submitted work; no financial relationships with any organizations that might have an interest in the submitted work in the previous three years; no other relationships or activities that could appear to have influenced the submitted work. Dr. Ahmad Masri received grants from Pfizer and Akcea (paid to OHSU) that is not related to this work.

## ACKNOWLEDGMENT

We thank Hospital Medicine staff, Intensive Care Unit (ICU) staff, Emergency Medicine staff, Advanced Practice Providers, nursing staff, medical assistants, and ancillary staff at Henry Ford West Bloomfield and Henry Ford Allegiance Hospitals, including: Anish Wadhwa, MD; Richard Santos, MD PhD; James Chauncey, MD; Rami Alzebdeh, MD; Yasser Aleech, MD; Imran Tarrar, MD; Sairia Dass, MD; Chidamber Alamelumagapuram, MD; Rahman Akhil, MD; Kamelia Albujoq, MD; Dominik Starosta, MD; Monika Grewal, MD; Angela Eke-Usim, MD; Amy Beaulac, MD; Tricia Stein, MD; Steven Rockoff, DO; Linoj Samuels, PhD; Beverly Duthie, NP; Victoria Churchill, NP; and Alex Wells, MPH.

We also thank Dr. Craig Hutchinson (Corizon Health) for his contribution. We also thank Dr. E. John Orav, PhD (Department of Biostatistics, Harvard University) and Dr. Heather J. Baer, ScD (Department of Epidemiology, Harvard University) for their support.

## CONTRIBUTORSHIP

Dr. Ahmed Altibi has contributed to the study design, study conduct, statistical analysis, and manuscript writing. Dr. Altibi is responsible for the overall content as guarantors and accepts full responsibility for the work and conduct to the study and has access to the data. The following authors have contributed significantly to the chart review and data collection process: Hassan Liaqat, Alexander A. Slota, Radhika Sheth, Lama Al Jebbawi, Matthew E. George, Allison LeDuc, Enas Abdallah, Luke R. Russel, Saniya Jain, Nariné Shirvanian. Drs Bhargava Pallavia, Ahmed Masri, and Vivek Kak have contributed to the study design, monitoring integrity of data, supervising the overall progress and quality of the study, and reviewed the manuscript of the study.

## Notes

### Competing Interest Statement

The authors have declared no competing interest.

### Funding Statement

All authors declare no support from any organization for the submitted work; no financial relationships with any organizations that might have an interest in the submitted work in the previous three years; no other relationships or activities that could appear to have influenced the submitted work.

### Author Declarations

The study was reviewed and approved by the Henry Ford Health System (HFHS) Institutional Review Board (IRB).

